# Coupling time-aware SNP thresholds with genetic markers to define bacterial transmission chains in hospital surveillance

**DOI:** 10.64898/2026.04.27.26351816

**Authors:** Judit Burgaya, Jelena Erdmann, Leonard Knegendorf, Claas Baier, Dmytro Strunin, Rasmus L. Marvig, Karen Leth Nielsen, Frederik Boetius Hertz, Dirk Schlüter, Susanne Häussler, Marco Galardini

**Author notes:** Shared last-authorship (Susanne Häussler, Marco Galardini).

## Abstract

Hospital settings can act as reservoirs for pathogens, with many nosocomial bacteria surviving for extended periods of time and spreading via contaminated environments, healthcare workers, or medical equipment. Infection prevention and control in hospital settings relies on accurately identifying transmission events to prevent further transmission. Whole-genome sequencing (WGS) offers the possibility to accurately identify bacterial strains and define transmission routes, yet traditional analyses often rely on fixed single nucleotide polymorphisms (SNP) thresholds that fail to account for the accumulation of variations over extended periods of time. We analyzed WGS surveillance data from bacterial isolates collected over five years in two hospitals in Germany and Denmark. By leveraging longitudinal sampling and repeated isolates from persistent infections, we estimated *in vivo* evolutionary rates from different strains belonging to three species: *Escherichia coli, Klebsiella pneumoniae* and *Pseudomonas aeruginosa*. Using species-level mean molecular clock rates, we developed a time-aware framework that defines transmission threshold based on expected SNP accumulation over time. Using this approach, transmission clusters were detected in 0.3% of *E. coli*, 9.3% of *K. pneumoniae* and 3.5% of *P. aeruginosa* isolates.

To understand the genetic factors underlying epidemic strain potential, we compared epidemic lineages part of transmission chains with sporadic lineages. We found that epidemic lineages of *K. pneumoniae* and *E. coli* had higher virulence scores than sporadic strains, with enrichment of siderophores and adhesion genes, while resistance scores were similar. Genome-wide association analyses revealed hundreds of variants associated with epidemic status, particularly in replication, recombination and repair mechanisms, as well as metabolism related functions. With the predicted virulence and resistance scores we could easily observe predicted phenotypic changes within transmission clusters, including the loss of virulence factors in an outbreak, and the emergence of resistance after antibiotic treatment.

## Introduction

Hospital environments act as persistent reservoirs for pathogens, increasing the risk of patient colonization and transmission^1^. Many nosocomial organisms can survive on surfaces for weeks to months, maintaining the potential to infect new hosts either through direct transmission via the patient’s contact with contaminated surfaces or indirect transmission via healthcare workers or medical equipment^2^. Selective pressures, including the host immune responses and antibiotic exposure, drive the fixation of mutations that enhance bacterial survival within the host. This rapid adaptation complicates infection control and contributes to the persistence and spread of high-risk clones^3–5^. Epidemic strains are genetically and phylogenetically distinct, and often associated with higher morbidity and mortality, thus requiring methods to accurately identify the transmission of these strains and subsequently implement evidence-based interventions. Understanding the emergence, evolution, and transmission of these strains is critical; however, large-scale comparative studies across multiple bacterial species remain limited.

Whole genome sequencing (WGS) allows for high-resolution comparison of pathogen genomes, making it possible to distinguish closely related isolates and to infer likely transmission events over short time scales. Pathogen genomic surveillance, or the sequencing of isolates irrespective of the presence of a suspected outbreak, has been increasingly adopted in routine microbiology and public health laboratories in recent years^6–8^. By integrating whole-genome data into ongoing surveillance rather than deploying it only reactively, such programmes provide the population-level context to distinguish sporadic cases from sustained transmission^9^. A number of prospective studies have already demonstrated the ability of WGS to identify pathogen transmission events that would otherwise be missed through standard epidemiological methods^10–12^. Depending on the bacterial species, WGS has also been found to have the potential to be a cost effective measure if coupled with interventions that are able to stop the identified transmission chains^13,14^.

The standard approach taken in genomic epidemiology relies on the use of predefined thresholds on the number of SNPs to identify samples that are part of a transmission event^15–22^. While this rather crude approach has helped in identifying *bona fide* pathogen transmission events, it has obvious limitations. One of them is the rather arbitrary choice of the threshold on the number of SNPs, which is then applied equally to all pairs of samples which might be closely related. However, if two pairs of samples differ in the amount of time that separates their isolation, it can be expected that the pair with the longer time span will have a greater number of SNPs separating them, potentially going over the predefined threshold. Recent studies have indeed tried to derive dynamic thresholds using a combination of empirical and modelling approaches^23,24^. The availability of ever larger WGS datasets, with the same clonal lineage being repeatedly sampled from the same patient, would allow for purely empirical approaches to be developed and tested for their ability to better identify transmission chains.

In this work, we leveraged a collection of whole-genome sequenced clinical isolates of bacteria belonging to three bacterial species (*E. coli, K. pneumoniae*, and *P. aeruginosa*) from two hospitals, and including patient and clinical metadata. The longitudinal sampling, over a 5-year period, included patients whose bacterial pathogen was sampled and sequenced multiple times over the course of the infection, allowing us to calibrate an empirical molecular clock, which we used to derive a dynamic SNP threshold. We used this approach to identify likely pathogen transmission chains, which allowed us to investigate the genetic determinants most associated with pathogen transmission. We also used simple virulence and resistance scores based on the presence of known genetic determinants, and used changes in these scores during transmission chains to identify instances of in-hospital adaptation of pathogens.

## Results

### Defining strain relatedness and putative transmission events using empirical molecular clock rates

We made use of a collection of 29,239 genomes from two tertiary care hospitals in Germany (Medizinische Hochschule Hannover, MHH) and Denmark (Rigshospitalet Copenhagen). For both hospitals, we had access to samples from three species: *E. coli, K. pneumoniae*, and *P. aeruginosa* with associated patient metadata (**Figure 1A-B**). The samples and respective genomic data and metadata spanned a period going between 2020 and 2024, with a rough average of 500 samples per month over the study period (**Supplementary Figure 1**). Crucially, a number of patients (N=52) had been sampled multiple times during the course of their hospital stay; this provided us with an opportunity to calculate an empirical rate of mutation accumulation and thus to calculate a dynamical SNP threshold that would change based on the time dividing pairs of samples.

**Figure 1.**
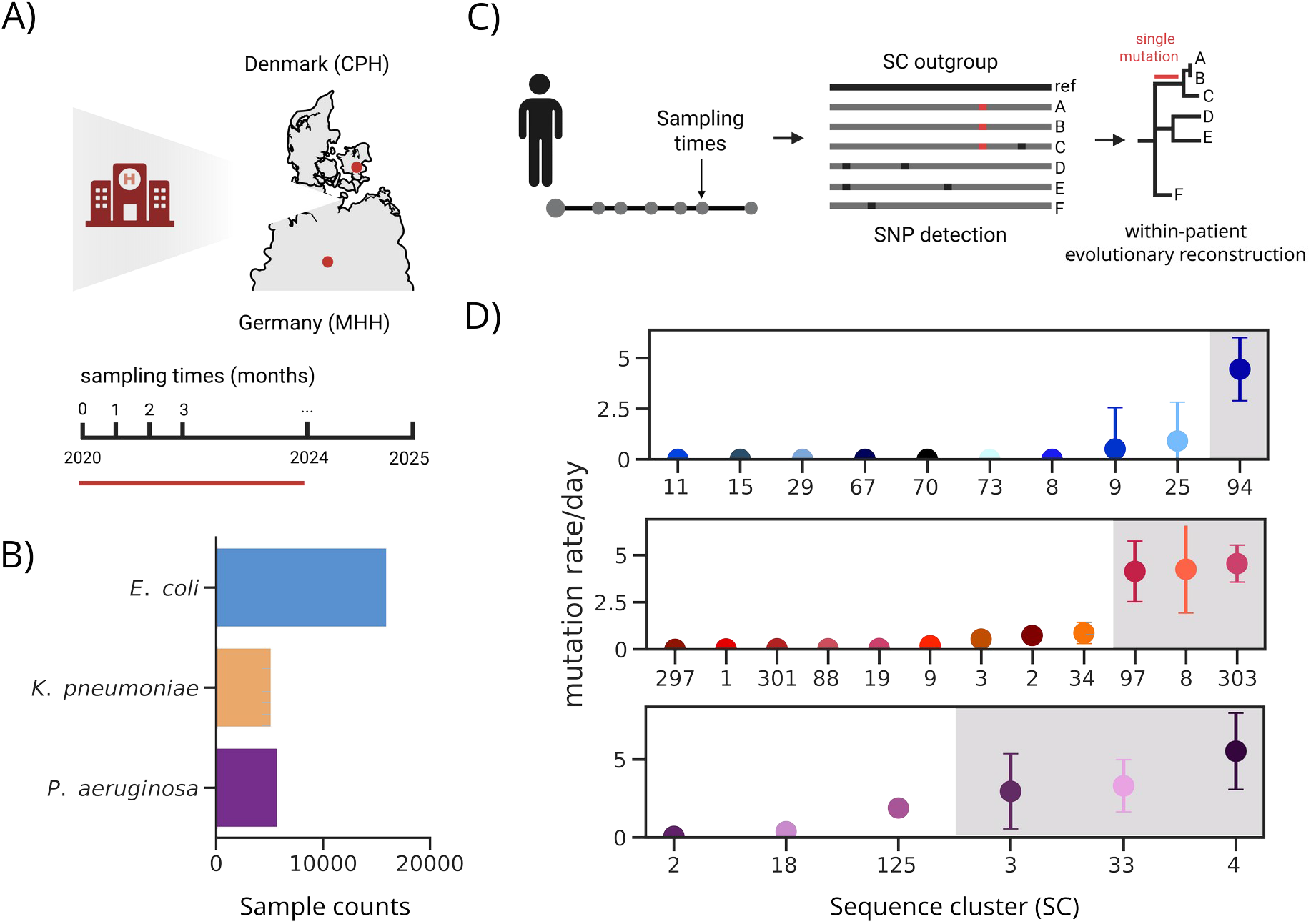
Study design and estimation of molecular clock rates. **A)** Genome collection assembled over a 5-year period from two tertiary-care hospitals in Germany (Medizinische Hochschule Hannover, MHH) and Denmark (Rigshospitalet, CPH). The line in red shows the time-span of samples used for the transmission analysis, while samples from 2024 were only used for the validation of the GWAS results. **B)** Analysis focused on three species with the largest available number of samples: *E. coli, K. pneumoniae*, and *P. aeruginosa*. **C)** Single-nucleotide polymorphisms (SNPs) were identified using an outgroup from the same sequence cluster (SC) to root within-patient phylogenies, and within-patient phylogenetic trees were constructed (Methods) **D)** Estimated mutation rates per day for *E. coli* (top), *K. pneumoniae* (middle), and *P. aeruginosa* (bottom) were obtained across different SCs and patients sampled at least 15 times over their hospital stay. Estimates within the grey shaded area correspond to hypermutator strains and were excluded to derive a conservativedynamic SNP threshold.

As a necessary first step we split all samples into sequence clusters (SCs), using a k-mer method. For each SC we obtained a reference-free alignment from which a phylogenetic tree was derived. We then obtained the SNP distances between all samples within each SC, which is a proxy for their relatedness. If two samples are part of a transmission chain, then the number of accumulated non-adaptive SNPs between them should be correlated with the time passed between their isolation, which can also be termed a “molecular clock”. For each of the three species, we calibrated this molecular clock using data from infected patients that were sampled at least 15 times over their hospital stay (52 patients with an average number of 21 samples each). For each patient we constructed a phylogenetic tree, incorporating an outgroup from the same SC that was sampled at an earlier time point, and when possible from a different patient (**Figure 1C**). We then used the sampling date to derive the rate of accumulation of SNPs for each patient.

We observed a separation in two regimes for the inferred molecular clock rates for the three species. For *K. pneumoniae*, nine of the patients showed consistent molecular clock rates, while three patients had substantially higher accumulation mutation rates (4-5 SNPs/day, **Figure 1D**). Similarly, for *E. coli* and *P. aeruginosa* one and three patients, respectively, also had higher mutation rates (4 SNPs/day and 3-6SNPs/day, **Figure 1D**). This elevated mutation rate could possibly reflect environmental pressures that induce a higher number of mutations or defects in DNA repair. For the SCs with increased mutation rate, we generally did not observe any significant differences in the mutational spectra across patients-SCs. We did however observe a slight increase in G/C transversions in the *P. aeruginosa* SC with elevated molecular clock rates, which might be linked to polymerase errors, and might suggest a selective advantage or adaptation under stress conditions, such as those observed in chronic infections (**Supplementary Figure 2**)^25,26^. We further mapped the identified variants and observed that the hypermutator *E. coli* strain, along with two of the three *K. pneumoniae* strains, harbored SNPs in *mut* genes. These genes play a major role in several DNA repair pathways and have been previously linked to hypermutator phenotypes in both experimental evolution studies and clinical isolates from chronic infections^27,28^. We therefore excluded these patients from the analysis in order to obtain a more conservative dynamic SNP threshold for each species. The molecular clock rates estimated from the remaining data were 0.3 SNPs/day for *K. pneumoniae*, 0.4 SNPs/day for *E. coli* and 0.7 SNPs/day for *P. aeruginosa*.

We used the inferred SNP accumulation rates to assess the genetic relatedness between all samples collected during the study period, and identify potential transmission chains (**Figure 2A**). Samples from the same SC that differed less than the expected number of SNPs given their difference in sampling date were marked as probable transmissions. In order to reduce false positives we did not consider as probable transmission samples that were more than 180 days distant, regardless of the number of SNPs between them. We first compared our approach, termed the 180d method, against the regularly used threshold of 20 SNPs, regardless of the intervening time between samples, termed the 20SNP method. For *E. coli* and *K. pneumoniae* we observed a similar proportion of samples participating in a putative transmission chain between the two methods, with 0.35% and 14.54% with 180d and 0.27% and 14.94% for the 20SNP. For *P. aeruginosa* the 180d method identified a higher proportion of potentially transmitting samples, 3.69% against 1.39% with 20SNP (**Figure 2B**). We calculated the distribution of SNP distances between samples in the same SC for the three species to further investigate these differences (**Figure 2C, Supplementary Figure 3**); while for *K. pneumoniae* the majority of samples had 10 or fewer SNPs dividing them, the distribution for *E. coli* and *P. aeruginosa* was more variable; we observed that the majority of samples had more than a 20 SNPs difference, with a peak of 120 and 170 SNPs for *E. coli* and *P. aeruginosa*, respectively. The difference in the number of inferred transmission events when using the dynamic threshold is then based on the higher inferred SNP accumulation rate for *P. aeruginosa* when compared to *E. coli*. Given that transmission chains persisting for very long periods of time (*e*.*g*. 10 years) have been described^29^, and given our estimate that *P. aeruginosa* accumulates 1 SNP every 1.4 days, our approach likely identifies bona-fide transmission chains.

**Figure 2.**
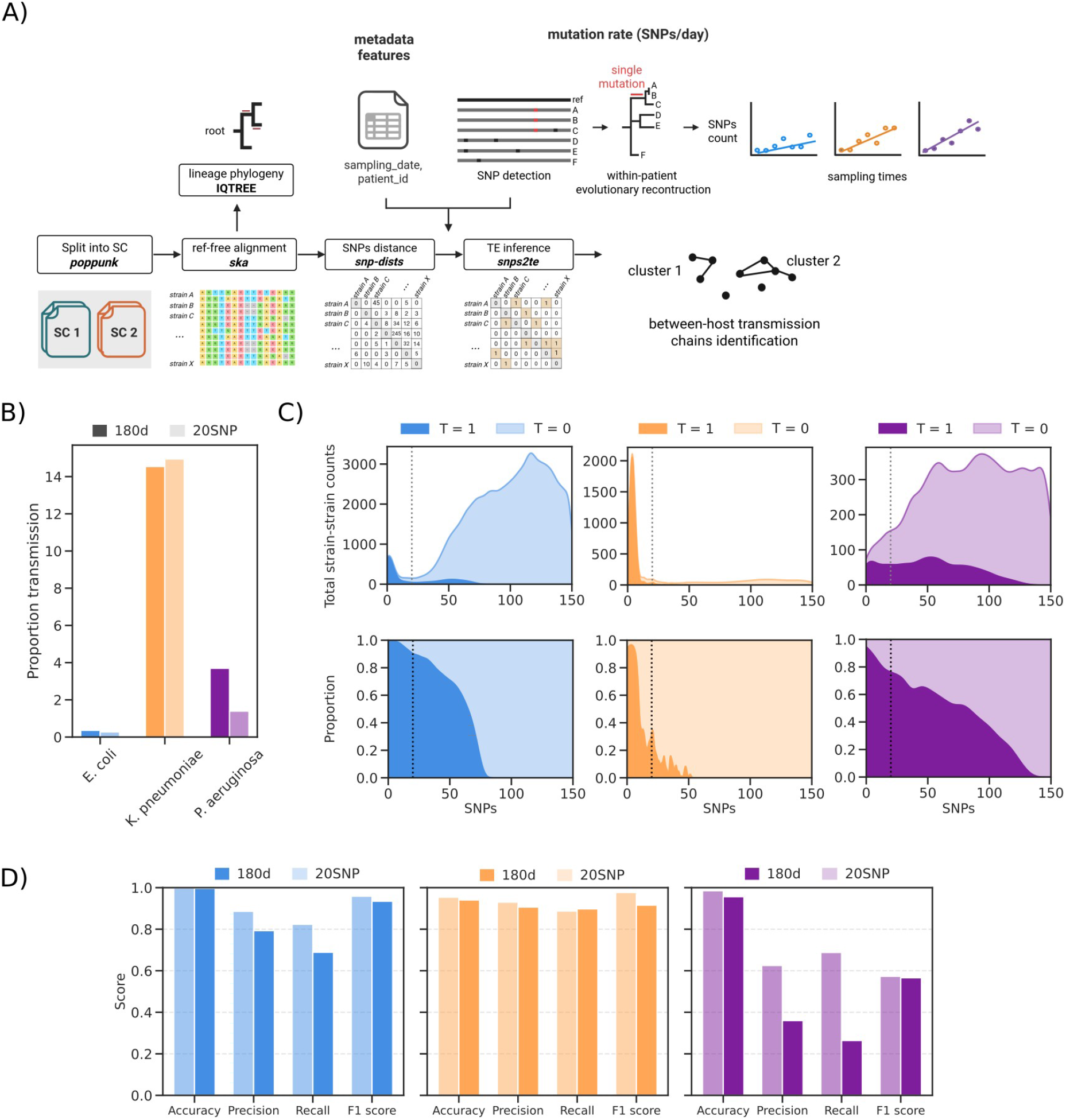
Identification of putative transmission events. **A)** Quantification of transmission events (Methods). **B)** Proportion of transmission events for *E. coli, K. pneumoniae* and *P. aeruginosa* estimated using a dynamic SNP threshold (180d) and a fixed threshold of 20 SNPs (20SNP). **C)** Distribution of the total number of identified putative transmission events (T=1) and the proportion of putative transmission events in relation to pairwise SNP distances for *E. coli* (blue), *K. pneumoniae* (orange) and *P. aeruginosa* (purple). **D)** Validation of the dynamic and fixed SNP threshold approaches using epidemiologically confirmed transmission events for each species.

We then used additional epidemiological information to validate the putative transmission events identified by both approaches. In particular, we used patient movement data, and identified epidemiologically confirmed transmissions when patients shared a ward (Methods), which represented our ground truth set for validation. We observed a similar value for the F1 score between the 20SNP and 180d methods for *K. pneumoniae*, with a higher precision for 20SNP and a higher recall for 180d. For the other two species we observed a similar F1 score between methods, but a lower value for precision and recall for 180d (**Figure 2D, Supplementary Figure 3**). These results might suggest that using a dynamic threshold might lead to a higher number of false positives, which however assumes that the epidemiological investigation identified all transmission events correctly.

### Genetic factors associated with epidemic strains

We next asked which features differentiated lineages that participated in a transmission chain (“epidemic”) from those that only caused isolated infections (“sporadic”). We first adopted a targeted approach, focused on factors related to virulence and AMR. We used sets of known virulence and antimicrobial resistance genes to derive a virulence and resistance score solely based on the genome sequences for the three species (**Supplementary Figure 4**). This scoring strategy had already been established for *K. pneumoniae*, and we developed a similar scoring scheme for *E. coli* and *P. aeruginosa* (**Methods**). We then compared these scores between epidemic and sporadic samples; we observed no differences across resistance scores between epidemic and sporadic strains across the three species (**Figure 3**). However, virulence differed between sporadic and epidemic strains for *K. pneumoniae* and *E. coli*. The majority of *K. pneumoniae* strains within transmission clusters encoded the aerobactin siderophore (*iuc*, virulence score 3), while most of the sporadic strains carried none (virulence score 0) or the yersiniabactin siderophore (*ybt*, virulence score 1). Most *E. coli* strains part of transmission clusters were associated with low-virulence factors related to colonization and adhesion (virulence score 1), while sporadic strains show also higher levels of ExPEC features (virulence score 4). We did not observe any difference across virulence patterns in *P. aeruginosa*, although as expected, we found that most strains were indicated as highly virulent with scores of 3 to 5, missing only a few of the known virulence factors (**Figure 3**).

**Figure 3.**
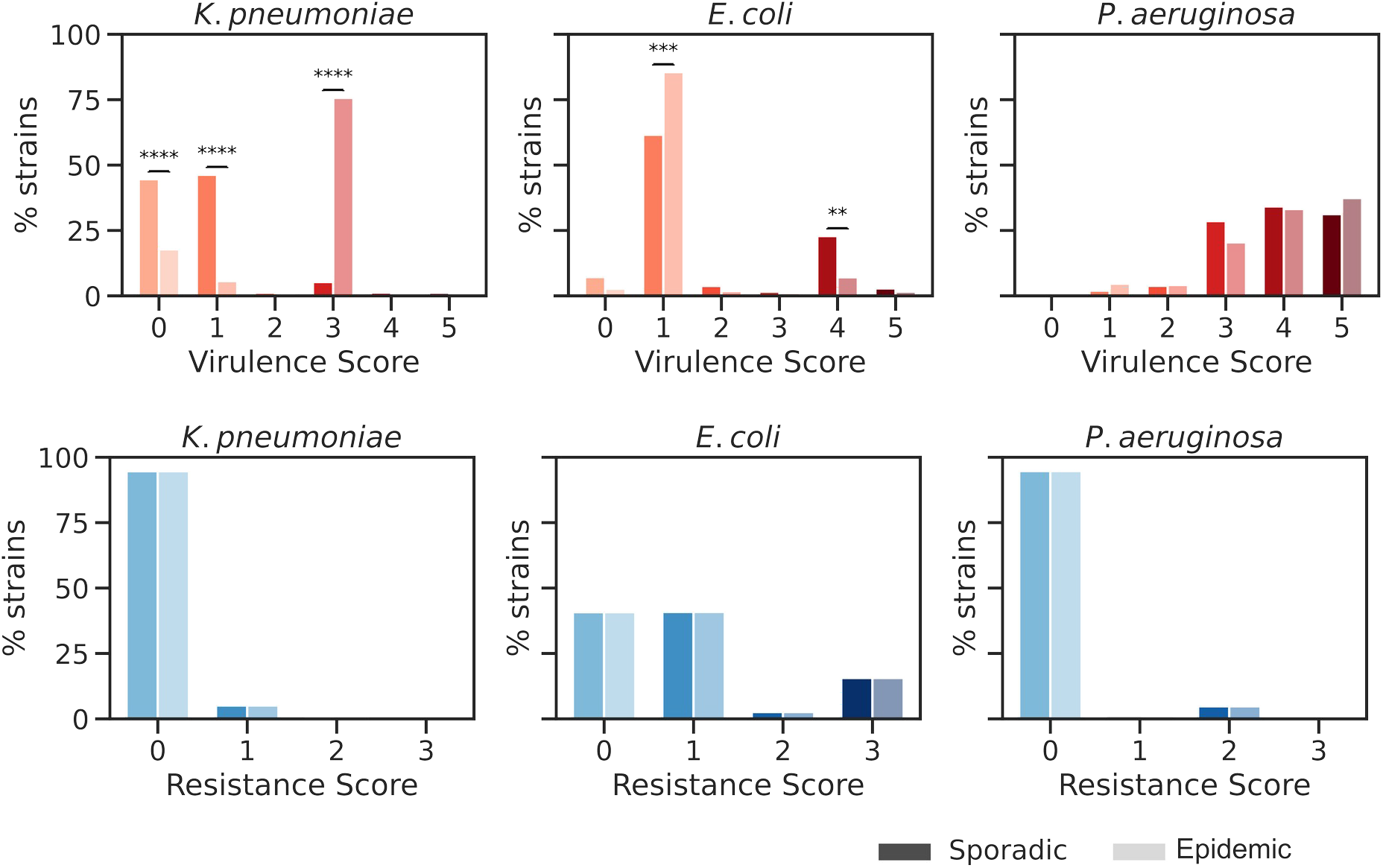
Virulence and resistance patterns across sporadic and epidemic strains. The percentage of strains within each virulence and resistance score was calculated, and significance was assessed using Fisher’s exact test.

We next used a hypothesis-free, untargeted approach to identify genetic variants associated with epidemic isolates. We first computed the narrow sense heritability (*h*^*2*^), or the fraction of phenotypic variance that is explained by additive genetic effects, which would give an indication of the presence of such variants in the genomes of the bacterial isolates. For all of the three species that we studied we observed a high heritability of the epidemic phenotype attributed to genetic variants: 76.27%, 89.09%, and 90.78% for *K. pneumoniae, E. coli* and *P. aeruginosa*, respectively. These high estimates were not fully due to broad population structure effects, as we computed much lower estimates for heritability when using SCs membership instead of variants: 27.12%, 19.89%, and 15.47% for *K. pneumoniae, E. coli* and *P. aeruginosa*, respectively (**Figure 4A-B**).

**Figure 4.**
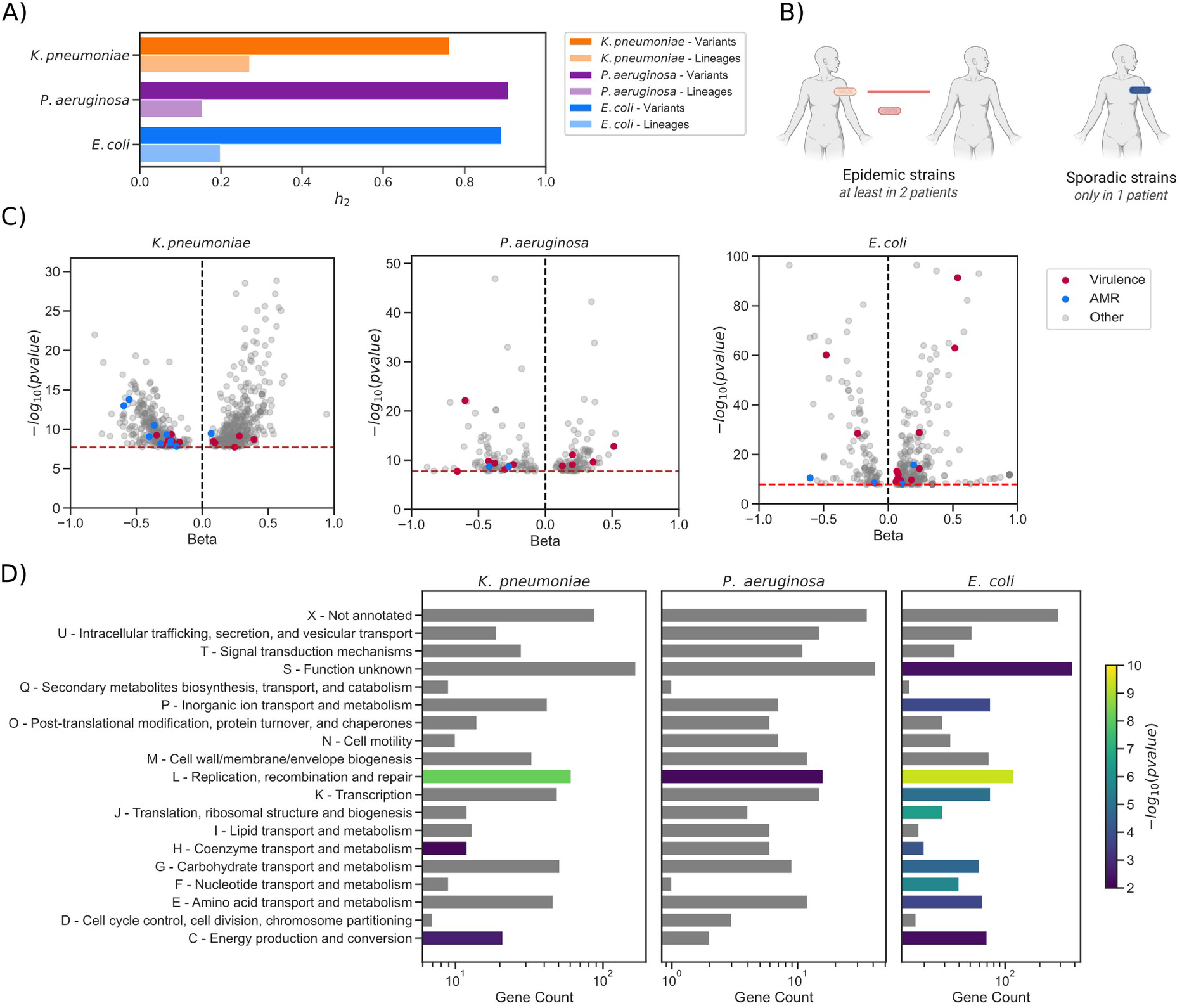
GWAS linking genetic variation to epidemic status. **A)** Variant- and lineage-level heritability estimates. **B)** Phenotype definition for association testing, with sporadic strains restricted to a single patient and epidemic strains shared among at least two patients. **C)**Genes associated with epidemic (positive beta) or sporadic (negative beta) strains. Only variants passing the significance threshold (red) are shown. Virulence- and AMR-related genes are indicated in red and blue, respectively. **D)** COG enrichment analysis of the associated variants.

We then run a genome-wide association study (GWAS) analysis, measuring the statistical association between the presence of genetic variants and the epidemic status of the isolates. In order to capture both core and accessory genome variation, we encoded genetic variants using k-mers. In order to partially correct for host factors we also included the sex and age of the infected patients as covariates. We used a linear mixed model (LMM) to run the associations, and then collated the variants passing the association threshold by the genes they mapped to. We identified a large number of genes associated with the epidemic or sporadic phenotype: 375, 1071, and 133 for *K. pneumoniae, E. coli*, and *P. aeruginosa*, respectively (**Figure 4C, Supplementary Figure 5**). Taken as a whole, we found the associated genes to be enriched in the L COG category (replication, recombination, and repair) across all species, and the H (energy production and conversion) and C (coenzyme transport and metabolism) COG categories for *K. pneumoniae* and *E. coli* (**Figure 4D**). We observed that a relatively large fraction of genes associated with either the epidemic or sporadic phenotype were annotated as virulence or AMR related genes: 12, 18, and 11 virulence genes and 10, 4, and 2 AMR genes for *K. pneumoniae, E. coli*, and *P. aeruginosa*, respectively (**Figure 4C**). Genes with variants associated with sporadic *K. pneumoniae* strains were enriched in adhesion-related genes, including *fimD/F* and *pilQ*^30^, while genes with variants associated with epidemic strains included toxins (*vapB/C*^31^ genes) and virulence plasmids associated genes (*virB*^32^). In *P. aeruginosa*, genes with variants associated with epidemic strains included genes linked to biofilm formation and adhesion, such as *epsE*^33^ and *psaB*^34^. For *E. coli*, genes with variants associated with epidemic strains included iron acquisition genes, including the *feoABC*^35^ operon and *yggX*^36^, alongside adhesion-related genes like *papC*^37^ and toxin genes such as *ccdA*^38^. For AMR genes, all three species’ sporadic strains included associated variants in *mdt* genes encoding for an efflux pump protein conferring antibiotic resistance^39^. Additionally, the sporadic strains of *K. pneumoniae* included variants in the *cmlA* gene conferring resistance to cloramphenicol^40^. In contrast, *K. pneumoniae* epidemic strains encoded associated variants in the *cusC* gene, which has been linked to tetracycline resistance^41^.

The presence of a relatively large number of variants associated with epidemic strains suggests that it might be possible to use genomic data to predict whether an isolate is more likely to be transmitted in a clinical environment. We therefore trained a ridge linear model trained on the presence of all genetic variants with a minor allele frequency (MAF) of at least 1%, again with host and sampling covariates added. We then validated the model’s ability to correctly identify epidemic isolates by running predictions on genomes isolated during 2024, which were not originally included in the study and therefore not used for model training. We observed that the model showed limited discriminatory power when applied to the independent dataset, indicating that genomic features are not sufficient to reliably distinguish between sporadic and epidemic strains (**Supplementary Figure 6**). These results suggest that, despite the presence of variants enriched among epidemic strains, their combined predictive signal does not generalize well to newly sampled isolates. This finding highlights that external factors, such as exposure to the pathogen or host comorbidities, may play an important role in shaping the transmission dynamics of bacterial strains, beyond the contribution of genomic variation alone.

### Changes in virulence and resistance scores during sustained transmission chains

As it has been previously demonstrated and as we have observed in this study, bacterial pathogens accumulate mutations during the course of transmission chains. Some of these mutations could affect infection-related phenotypes, leading to a potential impact on patient treatment and the risk of further transmission. To better understand this phenomenon from the perspective of infection research, we used the changes in virulence and resistance scores for epidemic strains. From the putative transmission chains with at least two patients on them, we observed that 9.4%, 18.8%, and 13.5% showed changes in the virulence score for *K. pneumoniae, E. coli* and *P. aeruginosa*, respectively, while 12.5%, 18.1%, and 9.6% of them presented changes in the resistance score.

Using this approach, we identified a clear change in the virulence score in isolates of *Klebsiella pneumoniae* SC34/ST86 (**Figure 5**). This SC is a major hyper-virulent *K. pneumoniae* (hvKP) lineage, typically associated with the K2 capsular polysaccharide, and is responsible for infections worldwide^42^. The hvKP strains are characterized by a virulence phenotype involving siderophores expression and a hypermucoid capsule, often encoded on mobile genetic elements. We identified a cluster of hvKP SC34/ST86 strains spanning a 4-month period in the Hannover hospital. The isolates did not encode for ESBLs or carbapenemases, but initially had a virulence score of 3. In four patients, however, we observed the loss of virulence-associated genes, resulting in a virulence score of 0. This reduction was driven by the loss of the *iuc* and *iro* loci, which encode the siderophores aerobactin and salmochelin, respectively, both of which are associated with invasive disease. Siderophores are essential for iron acquisition, facilitating bacterial growth and dissemination during systemic infections. Areobactin in particular has been shown to be a key determinant of hypervirulence in *K. pneumoniae*, while the salmochelin contributes to enhanced survival and persistence^43^. In addition to siderophore loss, the *rmpADC* operon, which regulates the hypermucoid phenotype and is associated with immune evasion, was also absent in these isolates^44^. Based on the likely origin of the contigs encoding these genes, they were likely located on an IncHI1B(pNDM-MAR) plasmid, which has been previously associated with invasive disease, which was absent in the same strains^45^.

**Figure 5.**
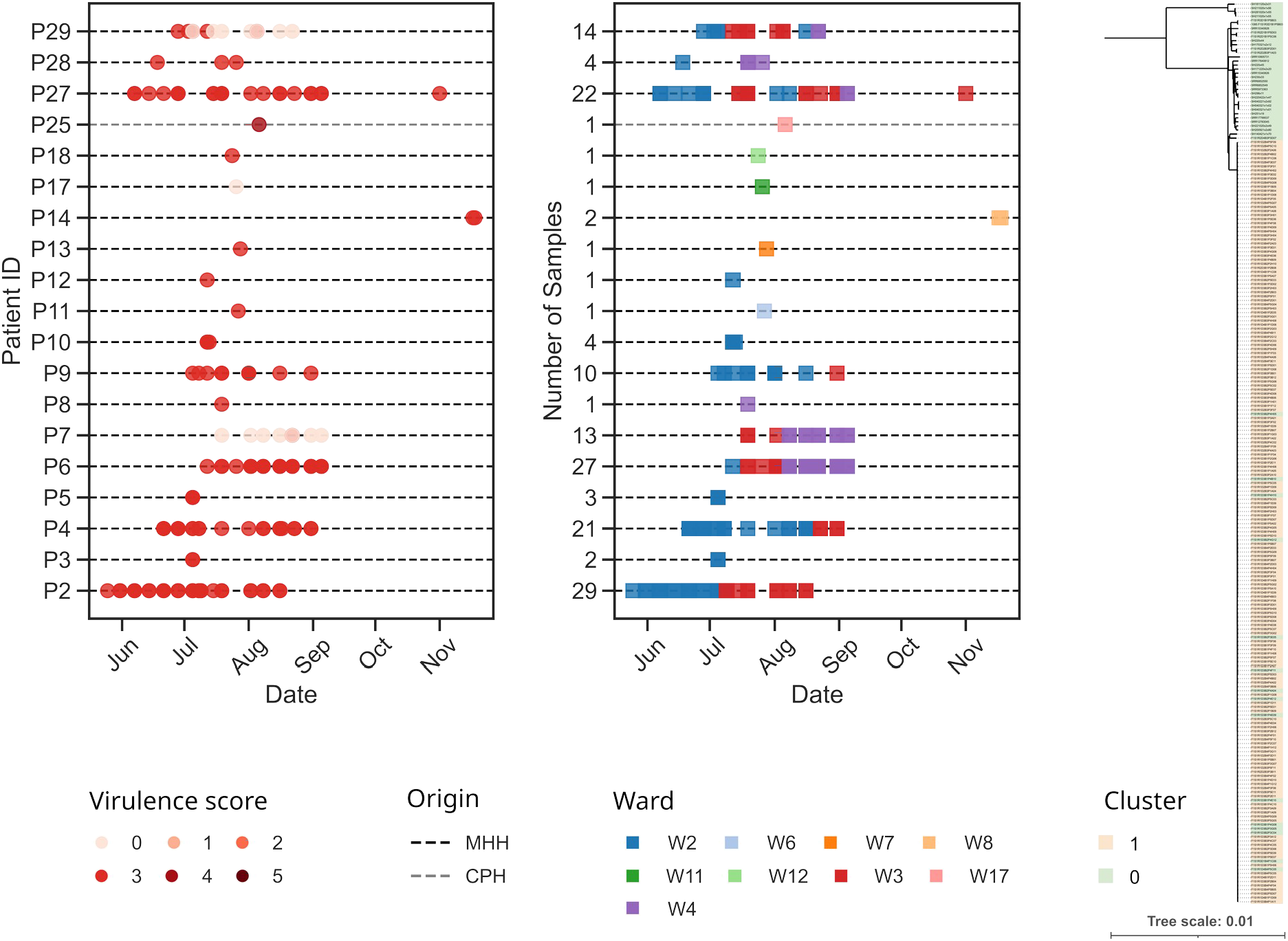
Distribution of *K. pneumoniae* SC34 samples by patient and hospital ward during the period in which the hvKP cluster was identified. The phylogenetic tree includes all *K. pneumoniae* SC34 strains, with cluster-associated isolates highlighted in orange.

The change in virulence score was first detected in the third sample of one patient (P29, **Figure 5**), obtained from a rectal swab. Earlier samples had been collected from nasopharynx and oropharynx swabs and both were carrying the virulent strain. Subsequent rectal swabs consistently lacked the virulence associated genes, and later nasopharyngeal and oropharyngeal samples showed the same phenotype. In one additional patient (P28), the decrease in virulence was also first detected in rectal swabs. In contrast, one patient (P7) appeared to have been initially colonized by the less virulent strain and later acquired the virulent strain. The final patient (P17) showed only sporadic colonization with the less virulent strain, detected in a urine sample, and no further sampling was performed. Notably, out of the 17, three patients had bloodstream infections, all caused by isolates carrying the virulence-associated genes. Previous studies have reported the loss of virulence factors as part of within-host adaptation^46^. Our observations are consistent with this phenomenon, particularly in isolates recovered from rectal swabs, which may reflect adaptation to intestinal colonization. At the same time, the presence of the virulence plasmid in bloodstream isolates highlights the importance of these genes in the development of invasive disease, suggesting a potential trade-off between efficient colonization and transmission versus the capacity to cause severe infections.

Similarly, we identified changes in the antimicrobial resistance score during transmission events. One example involved the transmission of a *K. pneumoniae* SC3/ST45 strain between two patients (**Figure 6**). In this case, the resistance score increased from 0 to 1 in one of the patients (P1), corresponding to the acquisition of the SHV-2 allele in the *bla*_SHV_ gene. This allele is characterized by a single amino acid substitution (Gly238Ser), which confers the *bla*_SHV_ gene an extended-spectrum β-lactamase (ESBL) activity^47,48^, including ceftriaxone^49^, which was indeed administered to patient P1 prior to the emergence of this allele. Patient P2 was infected with the same isolate, but in their case the resistance score did not increase in the three samples isolated during their hospital stay. Both patients were admitted to the same ward. Therefore, patient P2 may have acquired the strain from patient P1 through direct transmission, or alternatively, both patients may have acquired it from a common hospital source within the same ward.

**Figure 6.**
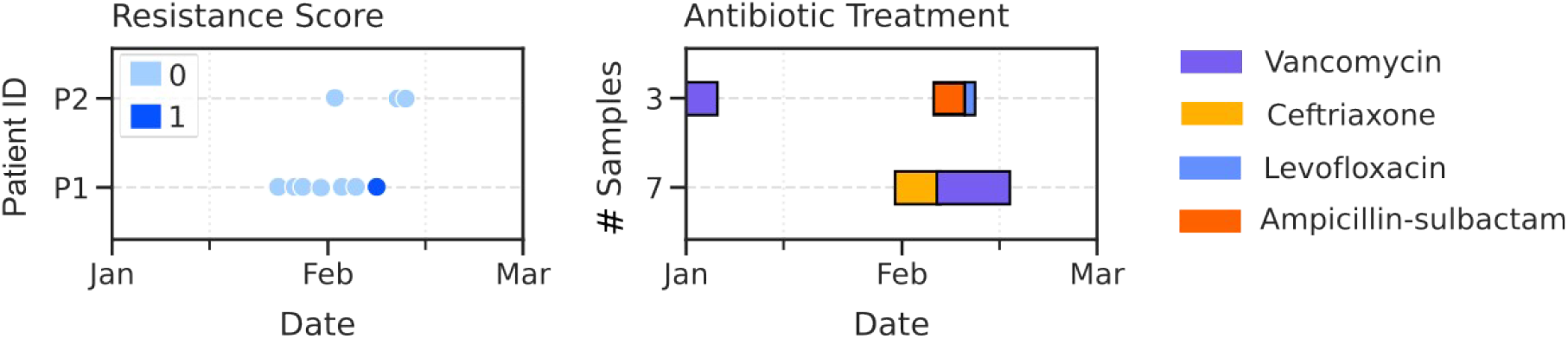
Distribution of *K. pneumoniae* SC3 samples by patient and antibiotic treatment during the transmission period. An increase in the resistance score from 0 to 1 is observed in one patient, corresponding to the acquisition of an ESBL. The right panel shows the antibiotic treatments administered to both patients during the same time window.

## Discussion

Infection prevention and control (IPC) in health-care settings has traditionally relied on a reactive response. Typically, samples are sequenced only after an outbreak has been identified, with the aim of reconstructing the most likely transmission routes. Daily surveillance monitors patients who test positive for pathogens of concern for nosocomial infection, and epidemiological investigations assess their connections in time and place, as well as any additional factors that might link cases. Although microbial typing may be used to define bacterial relatedness, conventional methods, such as MALDI-TOF, are generally thought to lack sufficient discriminatory power to resolve relatedness beyond the level of bacterial clones. Recent studies support the implementation of rapid response or prospective genomic epidemiology surveillance programmes, which enable earlier outbreak detection, thus potentially preventing infections from arising in the first place^10–14^.

Despite these clear benefits, the effectiveness of genomic surveillance ultimately depends on how accurately transmission clusters are defined. A central challenge lies in distinguishing true transmission events from unrelated cases; fixed SNP thresholds are commonly used to infer genomic relatedness, but these static cut-offs do not take bacterial evolution and time into account. Replication, within-host adaptation, and environmental persistence, are ongoing evolutionary processes that shape bacterial genomes with the continuous accumulation of variation across generations. Evidence from long-term evolution experiments and within-host evolution demonstrates that bacteria can accumulate mutations at different rates under different conditions and selective pressures^50–53^. We therefore sought to derive an empirical and dynamic SNP threshold from the same dataset to which it would be applied. We could do that thanks to the presence of patients that were repeatedly sampled during their hospital stay, which gave us confidence that all samples belonged to the same lineage and could thus be used as calibration. Compared with static SNP thresholds, this time-aware approach improves the identification of prolonged or intermittent transmission events while preserving epidemiological meaning. This is particularly relevant for opportunistic pathogens capable of persisting in hospital environments for extended periods of time^54,55^. Using this framework, we identified transmission clusters involving 9.3% of *Klebsiella pneumoniae*, 0.3% of *Escherichia coli*, and 3.5% of *Pseudomonas aeruginosa* isolates. Comparison with the fixed threshold showed broadly similar results for *K. pneumoniae* and *E. coli*, whereas the dynamic approach identified a larger number of transmission events for *P. aeruginosa*. Even though our approach was characterized by a lower concordance with epidemiological data, we believe that this “ground truth” set had the chief limitation of being limited to shared ward exposure between patients. Therefore it did not systematically account for shared medical equipment, procedures, or healthcare personnel. As a result, some transmission events may have been identified only as those with genetic links rather than epidemiological links, reflecting incomplete epidemiological data rather than true absence of contact. Previous studies have documented outbreaks linked to contaminated medical equipment in *P. aeruginosa*^56^ and to medical procedures involving *E. faecium*^57^, underscoring that genomically defined transmission clusters without an apparent ward connection may in fact represent overlooked transmission routes.

By computing virulence and resistance scores, we were able to rapidly identify putative phenotypic changes within transmission clusters. This framework provides a simple way to flag clinically relevant genomic changes that may influence treatment decisions or infection control strategies. While phenotypic antimicrobial susceptibility testing remains the clinical standard, genomics may provide a faster susceptibility estimate as point-of-care sequencing becomes available^58,59^. In contrast, changes affecting virulence cannot generally be measured with current clinical diagnostic tools. Several studies have demonstrated that specific virulence determinants are strongly associated with invasive disease, suggesting that monitoring virulence-associated genomic variation could help identify strains with an increased capacity to cause severe infections^60–62^.

Having identified a number of putative transmission events, the next question we asked was whether we could identify genetic determinants that favour transmission. For this purpose we applied a hypothesis-free approach, with which we identified a large number of variants associated with either epidemic or sporadic isolates across the three species. Many of these variants fall into functional categories related to replication, recombination and repair, as well as metabolic processes such as energy production and coenzyme metabolism. Several of the genes enriched in epidemic strains are involved in functions that plausibly contribute to transmission or persistence in clinical environments, including adhesion, biofilm formation, and iron acquisition, and may therefore represent targets for continued surveillance during sustained transmission events. Despite these associations, genomic features alone were insufficient to reliably predict whether a strain would behave as epidemic or sporadic when evaluated on an independent dataset. This limited predictive performance suggests that, although certain variants are enriched among epidemic isolates, transmission dynamics are likely shaped by a combination of bacterial genetic factors and external variables, including host susceptibility, patient contact networks, treatment regimes, and environmental exposure.

IPC of hospital-acquired infections is increasingly moving toward a proactive model based on whole-genome sequencing surveillance. While investing in preventative strategies could struggle to find institutional support because of its cost, the potential gains in DALYs and reduction in hospital stays demonstrates the value of early and targeted interventions. Effective implementation for whole-genome sequencing based surveillance requires the strategic selection of isolates and the integration with clinical metadata to prioritize clusters for investigation, which would further reduce cost and labour requirement. For example, in our dataset, the majority of infections are caused by only a few percent of *E. coli* clusters, suggesting that sequencing all *E. coli* may not be necessary. Instead, focusing on species associated with larger or more clinically significant outbreaks such as *P. aeruginosa* and *K. pneumoniae*, as well as *E. faecium, E. faecalis* or MRSA, may lead to greater “value for money” of genomic surveillance^13,63^. Developing a robust hospital-specific pipeline also depends on close collaboration between clinical teams, to ensure that turn-around times are sufficiently rapid to influence decision making. Our approach demonstrates that time-aware thresholds improve the detection of transmission clusters, allowing identification not only of short and acute outbreaks but also recurrent, hidden, or long-term pathogen circulation that may extend beyond the hospital environment. When combined with high-resolution clinical data, such an integrated surveillance framework has the potential to reduce infection rates, optimize IPC strategies, and improve both patient outcomes and resource utilization. Future work should focus on validating and optimizing these thresholds across diverse hospital settings, computing them for additional species of interest, assessing whether species- or lineage-specific thresholds are needed, to finally evaluate their impact in real-time genomic surveillance programs.

## Methods

### Isolates collection

We conducted a retrospective study on bacterial isolates from infected patients from two hospitals, the Hannover Medical School (MHH) in Hannover, Germany, and the Copenhagen University Hospital - Rigshospitalet (CPH) situated in Copenhagen, Denmark. The study involved the collection of WGS data from bacterial cultures derived from patient’s swabs for three bacterial species (*E. coli, K. pneumoniae*, and *P. aeruginosa*) and associated patient metadata. This included a total of 15,995, 5,286 and 5,962 samples, and 10,077, 2,862, and 2,862 unique patients for *E. coli, K. pneumoniae* and *P. aeruginosa*, respectively.

### Processing of bacterial genomic data

Illumina short-reads were obtained as previously described in Hertz *et al*.^64^ Assemblies and annotations were obtained using an in-house analysis pipeline that includes trimming with bbmap v36.49^65^, assembling the reads with shovill v1.0.4 using SPAdes v3.14^66^, and annotation with prokka v1.14.6^67^. Quality control (QC) was done based on all assemblies and annotations using QUAST v5.2^68^. The parameters used for the QC were the N50, contig length at 50% of the total genome assembly, the length of the genome, and the number of CDS on the annotations. The cut-off values were used according to each species (**Supplementary Table 1**), and were based on the distribution of the data and expected values for each species.

### Determination of population structure, pangenome, and virulence/resistance scores

The genomes were clustered into sequence clusters (SC) using PopPUNK v2.4^69^. The annotated assemblies were used as input for Panaroo v1.5^70^, which determined the core and accessory genes of the three species isolates. *Klebsiella pneumoniae* isolates were further annotated using Kleborate v2.3.2^71^. AbritAMR v1.0.14 was used to identify antimicrobial resistance and virulence determinants for the three species^72^. The tool further classifies the hits into resistance and virulence classes of interest, however it does not have a specific database for *P. aeruginosa* virulence genes. For this reason, we collected a conservative list of *Pseudomonas* species virulence factors obtained from the Virulence Factor Database (VFDB)^73^, Victors Virulence Factors (PHIDIAS)^74^ and curated by the Pseudomonas Genome Database (PseudoCAP)^75^ with focus being primarily on the reference strains PAO1 and PA14. We filtered the list for duplicated entries and classified the virulence genes into a category according to previously published data. The list of genes is available in **Supplementary Table 2** and in a code repository (https://github.com/jburgaya/virulence_pseudomonas_db).

We summarized the predicted AMR genes and VAGs into a virulence and resistance score. For *K. pneumoniae*, scores were defined as in Kleborate^71^. Similarly, resistance scores for *E. coli* and *P. aeruginosa* were adapted from the *K. pneumoniae* scheme, ranging from 0 to 3). Many strains carry multiple resistance genes across different drug classes, so a score of 0 does not exclude multidrug resistance (**Supplementary Table 3**). For *E. coli* the virulence score ranged from 0 to 5, and was based on factors associated with extra-intestinal pathogenicity. For *P*. aeruginosa, virulence was scored based on the absence of key virulence determinants, as most strains are inherently virulent. Virulence and resistance scores for all isolates are available in **Supplementary Table 3**.

### Estimating molecular clock rates

All patients with at least 15 bacterial samples each were selected for each species. An outgroup belonging to the same SC and from an earlier time point than the first sample was selected as the reference for SNP calling, when possible. In the absence of such an outgroup, the earliest isolate from the same patient was used as the reference. We called SNPs against the reference using snippy v4.6 with thresholds of 10 and 0.9 for the minimum coverage and minimum fraction of supporting reads, respectively^76^. We also constructed a phylogenetic tree for each patient using IQ-TREE v2.1.2, rooting each lineage to the reference isolate we selected^77^. In the absence of selection, the accumulation of SNPs occurs independently and at a relatively constant rate. To remove spurious SNPs originating from sequencing errors or mapping artifacts, we used a Poisson distribution approach to filter out outliers. We set an upper threshold based on the 95th percentile of the Poisson distribution and removed the samples with abnormal SNP values within this 5%. Finally, to infer the molecular clock rate, we performed linear regression of the root-to-tip distances for isolates from each time point using scipy v1.13.1^78^. The sampling dates and number of SNPs for each isolate used to estimate the molecular clock rates is available in **Supplementary Table 4**.

### Estimating transmission events

To estimate transmission events for each species, the assemblies were first split into SCs. For each SC, matching split *k-mer* sequences were aligned in a reference-free manner using ska v.0.3.7^79^. A lineage-specific phylogenetic tree was inferred using IQTREE v2.1.2, with the GTR+G+ASC model, and the GTR+G as an alternative model if the +ASC failed^77^. Single-nucleotide polymorphisms between strains were then counted from the alignment with snp-dists v0.8.2^80^. Transmission events were inferred by using the estimated molecular-clock with a 95% CI, based on the expected number of accumulated SNPs between two samples based on the distance between the two sampling dates. Samples with less than 10 SNPs were always defined as related, to account for really short sampling date differences. Samples separated by more than 180 days were never considered as transmission events. Samples coming from the same patient were de-duplicated. A transmission events network was built as:

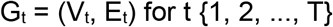

where:

G_t_:temporal network at each time step t (i.e. month).

V_t_: set of nodes (strains) present at time step t.

E_t_: set of directed edges representing inferred transmission between strains in Vt.

t: discrete time step corresponding to months (t=1, 2, …, T).

The pairwise SNP distances and inferred transmission events are available in **Supplementary Material 1**.

### Validation of transmission events identified by SNP distances

We validated the transmission events identified by our approach, termed 180d, as well as those identified by a fixed SNP distance threshold of 20 SNPs. We used hospital epidemiological data to define a set of confirmed transmission events, and used those as the “ground truth” for validation. An epidemiologically confirmed transmission was defined using multiple categories: *direct_impact* if patient1 was infected, patient2 was in the same ward on the same day, and patient2 tested positive for the same species within 14 days, *direct* if both patients were in the same ward on the same day, *indirect* if patient2 was in the same ward within 14 days as patient1, or *same_ward* if both patients were in the same ward at any time. Membership to any of these categories was considered an epidemiologically confirmed transmission. To quantify the agreement between genetic and epidemiological links, we computed the accuracy (proportion of genetic links that also have an epidemiological link), precision (proportion of true genetic links among the epidemiological links), recall (sensitivity - proportion of epidemiological links that were predicted as a genetic link) and F1-score (harmonic mean of precision and recall). These calculations were performed on patient movement data, which was available from the hospital journal system.

### Identification of genetic variants associated with transmission events

We used the microGWAS pipeline v0.3.0 to carry out the GWAS analysis ^81^., using default parameters. To facilitate the annotation of association hits, we included the following closed reference genomes from RefSeq, which were downloaded using the ncbi-genome-download package. We included the following genomes for E. coli: 536 (GCF_000013305.1), CFT073 (GCF_000007445.1), ED1a (GCF_000026305.1), IAI1 (GCF_000026265.1), IAI39 (GCF_000026345.1), MG1655 (GCF_000005845.2), UMN026 (GCF_000026325.1), and UTI89 (GCF_000013265.1). For K. pneumoniae we included a single reference: NTUH-K2044 (GCF_000009885.1). For P. aeruginosa we included two references: PAO1 (GCF_000006765.1) and UCBPP-PA14 (GCF_000014625.1). The annotated list of associated genes for each species is available in **Supplementary Material 2**. We used unitig-caller v1.3.1^82^ to make variant calls in the test population, and the elastic net regularization ridge model previously trained using pyseer v1.3.6^83,84^ to predict the transmission phenotype in the 2023-2024 data.

### Identification of putative phenotypic changes during hospital transmission

We investigated whether transmission chains were associated with changes in virulence or resistance by using the different scores. For isolates belonging to the same transmission cluster, we compared virulence and resistance scores across patients and sampling time points. For a subset of clusters, information on antibiotic treatment was manually retrieved from the hospital information system. Although not available for all patients, these data allowed us to contextualize some resistance changes and whether the acquisition of resistance determinants occurred as a result of the selective pressure induced during antimicrobial therapy.

## Data Availability

The collection of patient data was approved by the respective ethical committees from both hospitals. In order to protect the privacy of the participating patients, their metadata cannot be provided publicly. All code and scripts for analysis and figures are available as a code repository on GitHub (https://github.com/jburgaya/2025_genomics_transmission).

## Author contributions

MG, SH: conceptualization, funding acquisition. SH, KLN, RLM, CB, LK, DmS, DS, FBH: resources. JE, DmS: data curation. JB, MG: methodology, writing – original draft. JB: investigation, visualization. MG: supervision. All authors: writing – review & editing.

## Acknowledgements

This work was supported by the Deutsche Forschungsgemeinschaft (DFG, German Research Foundation) under Germany’s Excellence Strategy – EXC 2155 (Project No. 390874280). J.B. was supported by the Hannover Biomedical Research School (HBRS) and the Centre for Infection Biology (ZIB). JB was further supported by the Deutsche Forschungsgemeinschaft grant number GA 3191/1-1. S.H. received funding within the SFB/TRR-298-SIIRI—Project-ID 426335750 and in the SPP2389 (HA 3299/9–1, AOBJ: 687646), and from the Novo Nordisk Foundation (NNF 18OC0033946).

## References

1. Kanamori, H., Rutala, W. A., Sickbert-Bennett, E. E. & Weber, D. J. Role of the contaminated environment in transmission of multidrug-resistant organisms in nursing homes and infection prevention. Am. J. Infect. Control 51, A151–A157 (2023).

2. Kramer, A. et al. How long do bacteria, fungi, protozoa, and viruses retain their replication capacity on inanimate surfaces? A systematic review examining environmental resilience versus healthcare-associated infection risk by “fomite-borne risk assessment”. Clin. Microbiol. Rev. 37, e00186–23 (2024).

3. Pastor-Satorras, R. & Vespignani, A. Epidemic dynamics and endemic states in complex networks. Phys. Rev. E 63, 066117 (2001).

4. Lloyd-Smith, J. O., Schreiber, S. J., Kopp, P. E. & Getz, W. M. Superspreading and the effect of individual variation on disease emergence. Nature 438, 355–359 (2005).

5. Morens, D. M., Folkers, G. K. & Fauci, A. S. The challenge of emerging and re-emerging infectious diseases. Nature 430, 242–249 (2004).

6. Aanensen, D. M. et al. Whole-Genome Sequencing for Routine Pathogen Surveillance in Public Health: a Population Snapshot of Invasive Staphylococcus aureus in Europe. mBio 7, e00444–16 (2016).

7. Sundermann, A. J. et al. Whole-Genome Sequencing Surveillance and Machine Learning of the Electronic Health Record for Enhanced Healthcare Outbreak Detection. Clin. Infect. Dis. 75, 476–482 (2022).

8. Peacock, S. J., Parkhill, J. & Brown, N. M. Changing the paradigm for hospital outbreak detection by leading with genomic surveillance of nosocomial pathogens. Microbiology 164, 1213–1219 (2018).

9. Sundermann, A. J. et al. Pathogen genomics in healthcare: overcoming barriers to proactive surveillance. Antimicrob. Agents Chemother. 69, e01479–24 (2025).

10. Sherry, N. L. et al. Multi-site implementation of whole genome sequencing for hospital infection control: A prospective genomic epidemiological analysis. Lancet Reg. Health – West. Pac. 23, (2022).

11. Forde, B. M. et al. Clinical Implementation of Routine Whole-genome Sequencing for Hospital Infection Control of Multi-drug Resistant Pathogens. Clin. Infect. Dis. 76, e1277–e1284 (2023).

12. Sobkowiak, A. et al. The dark matter of bacterial genomic surveillance—antimicrobial resistance plasmid transmissions in the hospital setting. J. Clin. Microbiol. 63, e00121–25 (2025).

13. Hertz, F. B. et al. Estimating the potential economic and health impact of integrated genomic surveillance in a hospital setting. Clin. Microbiol. Infect. https://doi.org/10.1016/j.cmi.2025.09.021 (2025) doi:10.1016/j.cmi.2025.09.021.

14. Waggle, K. D. et al. Methods for cost-efficient, whole genome sequencing surveillance for enhanced detection of outbreaks in a hospital setting. 2024.02.16.24302955 Preprint at 10.1101/2024.02.16.24302955 (2025).

15. Raven, K. E. et al. Complex Routes of Nosocomial Vancomycin-Resistant Enterococcus faecium Transmission Revealed by Genome Sequencing. Clin. Infect. Dis. 64, 886–893 (2017).

16. Berbel Caban, A. et al. PathoSPOT genomic epidemiology reveals under-the-radar nosocomial outbreaks. Genome Med. 12, 96 (2020).

17. Marmor, A., Daveson, K., Harley, D., Coatsworth, N. & Kennedy, K. Two carbapenemase-producing Enterobacteriaceae outbreaks detected retrospectively by whole-genome sequencing at an Australian tertiary hospital. Infect. Dis. Health 25, 30–33 (2020).

18. Sundermann, A. J. et al. Outbreak of Pseudomonas aeruginosa Infections from a Contaminated Gastroscope Detected by Whole Genome Sequencing Surveillance. Clin. Infect. Dis. 73, e638–e642 (2021).

19. Sundermann, A. J. et al. Outbreak of Vancomycin-resistant Enterococcus faecium in Interventional Radiology: Detection Through Whole-genome Sequencing-based Surveillance. Clin. Infect. Dis. 70, 2336–2343 (2020).

20. Jakharia, K. K. et al. Use of whole-genome sequencing to guide a Clostridioides difficile diagnostic stewardship program. Infect. Control Hosp. Epidemiol. 40, 804–806 (2019).

21. Gona, F. et al. Comparison of core-genome MLST, coreSNP and PFGE methods for Klebsiella pneumoniae cluster analysis. Microb. Genomics 6, (2020).

22. Rose, R. et al. Molecular surveillance of methicillin-resistant Staphylococcus aureus genomes in hospital unexpectedly reveals discordance between temporal and genetic clustering. Am. J. Infect. Control 49, 59–64 (2021).

23. Stimson, J. et al. Beyond the SNP Threshold: Identifying Outbreak Clusters Using Inferred Transmissions. Mol. Biol. Evol. 36, 587–603 (2019).

24. Duval, A., Opatowski, L. & Brisse, S. Defining genomic epidemiology thresholds for common-source bacterial outbreaks: a modelling study. Lancet Microbe 4, e349–e357 (2023).

25. Khil, P. P. et al. Dynamic Emergence of Mismatch Repair Deficiency Facilitates Rapid Evolution of Ceftazidime-Avibactam Resistance in Pseudomonas aeruginosa Acute Infection. mBio 10, e01822–19 (2019).

26. da Cruz Nizer, W.S. et al. Oxidative Stress Response in Pseudomonas aeruginosa. Pathog. Basel Switz. 10, 1187 (2021).

27. Hall, K. M., Pursell, Z. F. & Morici, L. A. The role of the Pseudomonas aeruginosa hypermutator phenotype on the shift from acute to chronic virulence during respiratory infection. Front. Cell. Infect. Microbiol. 12, 943346 (2022).

28. Couce, A. et al. Mutator genomes decay, despite sustained fitness gains, in a long-term experiment with bacteria. Proc. Natl. Acad. Sci. 114, (2017).

29. Hoffmann, M. et al. Temporal Dynamics of Salmonella enterica subsp. enterica Serovar Agona Isolates From a Recurrent Multistate Outbreak. Front. Microbiol. 11, (2020).

30. Lillington, J., Geibel, S. & Waksman, G. Biogenesis and adhesion of type 1 and P pili. Biochim. Biophys. Acta BBA - Gen. Subj. 1840, 2783–2793 (2014).

31. Zhang, Y. X. et al. Characterization of a novel toxin-antitoxin module, VapBC, encoded by Leptospira interrogans chromosome. Cell Res. 14, 208–216 (2004).

32. Socea, J. N., Bowman, G. R. & Wing, H. J. VirB, a Key Transcriptional Regulator of Virulence Plasmid Genes in Shigella flexneri, Forms DNA-Binding Site-Dependent Foci in the Bacterial Cytoplasm. J. Bacteriol. 203, (2021).

33. Guttenplan, S. B., Blair, K. M. & Kearns, D. B. The EpsE Flagellar Clutch Is Bifunctional and Synergizes with EPS Biosynthesis to Promote Bacillus subtilis Biofilm Formation. PLoS Genet. 6, e1001243 (2010).

34. Soto, G. E. & Hultgren, S. J. Bacterial Adhesins: Common Themes and Variations in Architecture and Assembly. J. Bacteriol. 181, 1059–1071 (1999).

35. Lau, C. K. Y., Krewulak, K. D. & Vogel, H. J. Bacterial ferrous iron transport: the Feo system. FEMS Microbiol. Rev. 40, 273–298 (2016).

36. Osborne, M. J., Siddiqui, N., Landgraf, D., Pomposiello, P. J. & Gehring, K. The solution structure of the oxidative stress-related protein YggX from Escherichia coli. Protein Sci. 14, 1673–1678 (2005).

37. Norgren, M., Bága, M., Tennent, J. M. & Normark, S. Nucleotide sequence, regulation and functional analysis of the papC gene required for cell surface localization of Pap pili of uropathogenic Escherichia coli. Mol. Microbiol. 1, 169–178 (1987).

38. Chandra, S. et al. The High Mutational Sensitivity of ccdA Antitoxin Is Linked to Codon Optimality. Mol. Biol. Evol. 39, msac187 (2022).

39. Perreten, V., Schwarz, F. V., Teuber, M. & Levy, S. B. Mdt(A), a new efflux protein conferring multiple antibiotic resistance in Lactococcus lactis and Escherichia coli. Antimicrob. Agents Chemother. 45, 1109–1114 (2001).

40. Kuang, Q. et al. Detection and characterization of heteroresistance to chloramphenicol in Klebsiella pneumoniae isolates. BMC Microbiol. 25, 582 (2025).

41. Chen, D. et al. CusS-CusR Two-Component System Mediates Tigecycline Resistance in Carbapenem-Resistant Klebsiella pneumoniae. Front. Microbiol. 10, 3159 (2020).

42. Zhang, Y. et al. First report of two rapid-onset fatal infections caused by a newly emerging hypervirulent K. Pneumonia ST86 strain of serotype K2 in China. Front. Microbiol. 6, (2015).

43. Lam, M. M. C. et al. Tracking key virulence loci encoding aerobactin and salmochelin siderophore synthesis in Klebsiella pneumoniae. Genome Med. 10, 77 (2018).

44. Salisbury, S. M. et al. The acquisition of rmpADS can increase virulence of classical Klebsiella pneumoniae in the absence of other hypervirulence-associated genes. Preprint at 10.1101/2025.09.20.677538 (2025).

45. Spadar, A., Perdigão, J., Campino, S. & Clark, T. G. Genomic analysis of hypervirulent Klebsiella pneumoniae reveals potential genetic markers for differentiation from classical strains. Sci. Rep. 12, 13671 (2022).

46. Zaborskytė, G., Hjort, K., Lytsy, B. & Sandegren, L. Parallel within-host evolution alters virulence factors in an opportunistic Klebsiella pneumoniae during a hospital outbreak. Nat. Commun. 16, 8727 (2025).

47. Lee, K.-Y., Hopkins, J. D., Syvanen, M. & O’Brien, T. F. Gly-238-Ser substitution changes the substrate specificity of the SHV class A β-lactamases. Proteins Struct. Funct. Bioinforma. 11, 45–51 (1991).

48. Tsang, K. K. et al. Diversity, functional classification and genotyping of SHV β-lactamases in Klebsiella pneumoniae. Microb. Genomics 10, 001294 (2024).

49. Randegger, C. C., Keller, A., Irla, M., Wada, A. & Hächler, H. Contribution of Natural Amino Acid Substitutions in SHV Extended-Spectrum β-Lactamases to Resistance against Various β-Lactams. Antimicrob. Agents Chemother. 44, 2759–2763 (2000).

50. Tenaillon, O. et al. Tempo and mode of genome evolution in a 50,000-generation experiment. Nature 536, 165–170 (2016).

51. Wiser, M. J., Ribeck, N. & Lenski, R. E. Long-Term Dynamics of Adaptation in Asexual Populations. Science 342, 1364–1367 (2013).

52. La Rosa, R., Rossi, E., Feist, A. M., Johansen, H. K. & Molin, S. Compensatory evolution of Pseudomonas aeruginosa’s slow growth phenotype suggests mechanisms of adaptation in cystic fibrosis. Nat. Commun. 12, 3186 (2021).

53. Chomkatekaew, C. et al. A 28-year evolution experiment on Burkholderia pseudomallei survival in nutrient-depleted sterile water. 2026.02.06.704356 Preprint at 10.64898/2026.02.06.704356 (2026).

54. Kanamori, H., Rutala, W. A., Sickbert-Bennett, E. E. & Weber, D. J. Role of the contaminated environment in transmission of multidrug-resistant organisms in nursing homes and infection prevention. Am. J. Infect. Control 51, A151–A157 (2023).

55. Bourdin, T. et al. Ecological dynamics of three persistent opportunistic pathogens in hospital sinks and their potential antagonistic bacteria. mSystems 0, e01546–25 (2026).

56. Sundermann, A. J. et al. Outbreak of Pseudomonas aeruginosa Infections from a Contaminated Gastroscope Detected by Whole Genome Sequencing Surveillance. Clin. Infect. Dis. 73, e638–e642 (2021).

57. Sundermann, A. J. et al. Outbreak of Vancomycin-resistant Enterococcus faecium in Interventional Radiology: Detection Through Whole-genome Sequencing-based Surveillance. Clin. Infect. Dis. 70, 2336–2343 (2020).

58. Břinda, K. et al. Rapid inference of antibiotic resistance and susceptibility by genomic neighbour typing. Nat. Microbiol. 5, 455–464 (2020).

59. Sauerborn, E. et al. Detection of hidden antibiotic resistance through real-time genomics. Nat. Commun. 15, 5494 (2024).

60. Lees, J. A. et al. Joint sequencing of human and pathogen genomes reveals the genetics of pneumococcal meningitis. Nat. Commun. 10, 2176 (2019).

61. Chaguza, C. et al. Bacterial genome-wide association study of hyper-virulent pneumococcal serotype 1 identifies genetic variation associated with neurotropism. Commun. Biol. 3, 559 (2020).

62. Burgaya, J. et al. The bacterial genetic determinants of Escherichia coli capacity to cause bloodstream infections in humans. PLOS Genet. 19, e1010842 (2023).

63. Sundermann, A. J. et al. Whole-Genome Sequencing Surveillance and Machine Learning of the Electronic Health Record for Enhanced Healthcare Outbreak Detection. Clin. Infect. Dis. 75, 476–482 (2022).

64. Hertz, F. B. et al. Evaluating the economic and health impact of proactive genomic epidemiology in a hospital setting. Preprint at 10.1101/2025.01.30.25321399 (2025).

65. Bushnell, B. BBMap: A Fast, Accurate, Splice-Aware Aligner. (2014).

66. Bankevich, A. et al. SPAdes: A New Genome Assembly Algorithm and Its Applications to Single-Cell Sequencing. J. Comput. Biol. 19, 455–477 (2012).

67. Seemann, T. Prokka: rapid prokaryotic genome annotation. Bioinformatics 30, 2068–2069 (2014).

68. Gurevich, A., Saveliev, V., Vyahhi, N. & Tesler, G. QUAST: quality assessment tool for genome assemblies. Bioinformatics 29, 1072–1075 (2013).

69. Lees, J. A. et al. Fast and flexible bacterial genomic epidemiology with PopPUNK. Genome Res. 29, 304–316 (2019).

70. Tonkin-Hill, G. et al. Producing polished prokaryotic pangenomes with the Panaroo pipeline. Genome Biol. 21, 180 (2020).

71. Lam, M. M. C. et al. A genomic surveillance framework and genotyping tool for Klebsiella pneumoniae and its related species complex. Nat. Commun. 12, 4188 (2021).

72. Sherry, N. L. et al. An ISO-certified genomics workflow for identification and surveillance of antimicrobial resistance. Nat. Commun. 14, 60 (2023).

73. Chen, L. VFDB: a reference database for bacterial virulence factors. Nucleic Acids Res. 33, D325–D328 (2004).

74. Sayers, S. et al. Victors: a web-based knowledge base of virulence factors in human and animal pathogens. Nucleic Acids Res. 47, D693–D700 (2019).

75. Winsor, G. L. Pseudomonas aeruginosa Genome Database and PseudoCAP: facilitating community-based, continually updated, genome annotation. Nucleic Acids Res. 33, D338–D343 (2004).

76. Torster Seemann snippy: fast bacterial variant calling from NGS reads.

77. Minh, B.Q. et al. IQ-TREE 2: New Models and Efficient Methods for Phylogenetic Inference in the Genomic Era. Mol. Biol. Evol. 37, 1530–1534 (2020).

78. Virtanen, P. et al. SciPy 1.0: fundamental algorithms for scientific computing in Python. Nat. Methods 17, 261–272 (2020).

79. Derelle, R. et al. Seamless, rapid, and accurate analyses of outbreak genomic data using split k -mer analysis. Genome Res. 34, 1661–1673 (2024).

80. Seemann, T., Klötzl, F. & J Page, A. snp-dists.

81. Burgaya, J., Damaris, B.F., Fiebig, J. & Galardini, M. microGWAS: a computational pipeline to perform large-scale bacterial genome-wide association studies. Microb. Genomics 11, (2025).

82. Holley, G. & Melsted, P. Bifrost – Highly parallel construction and indexing of colored and compacted de Bruijn graphs. Preprint at 10.1101/695338 (2019).

83. Lees, J. A., Galardini, M., Bentley, S. D., Weiser, J. N. & Corander, J. pyseer: a comprehensive tool for microbial pangenome-wide association studies. Bioinformatics 34, 4310–4312 (2018).

84. Lees, J. A., Tien Mai, T., Galardini, M., Wheeler, N. E. & Corander, J. Improved inference and prediction of bacterial genotype-phenotype associations using pangenome-spanning regressions. Preprint at 10.1101/852426 (2019).

