## Supplementary Materials for "Coupling time-aware SNP thresholds with genetic markers to define bacterial transmission chains in hospital surveillance"

### Supplementary Tables

**Supplementary Table 1:** annotated assemblies QC cutoffs for each species
**Supplementary Table 2:** list of *P. aeruginosa* virulence genes
**Supplementary Table 3:** list of virulence and resistance scores for all isolates
**Supplementary Table 4:** number of SNPs and sampling dates for the isolates used for the calibration of the molecular clocks

### Supplementary Materials

**Supplementary Material 1:** pairwise SNP distances and inferred transmission events
**Supplementary Material 2:** annotated list of genes associated with transmission status for the three species

### Supplementary Figures

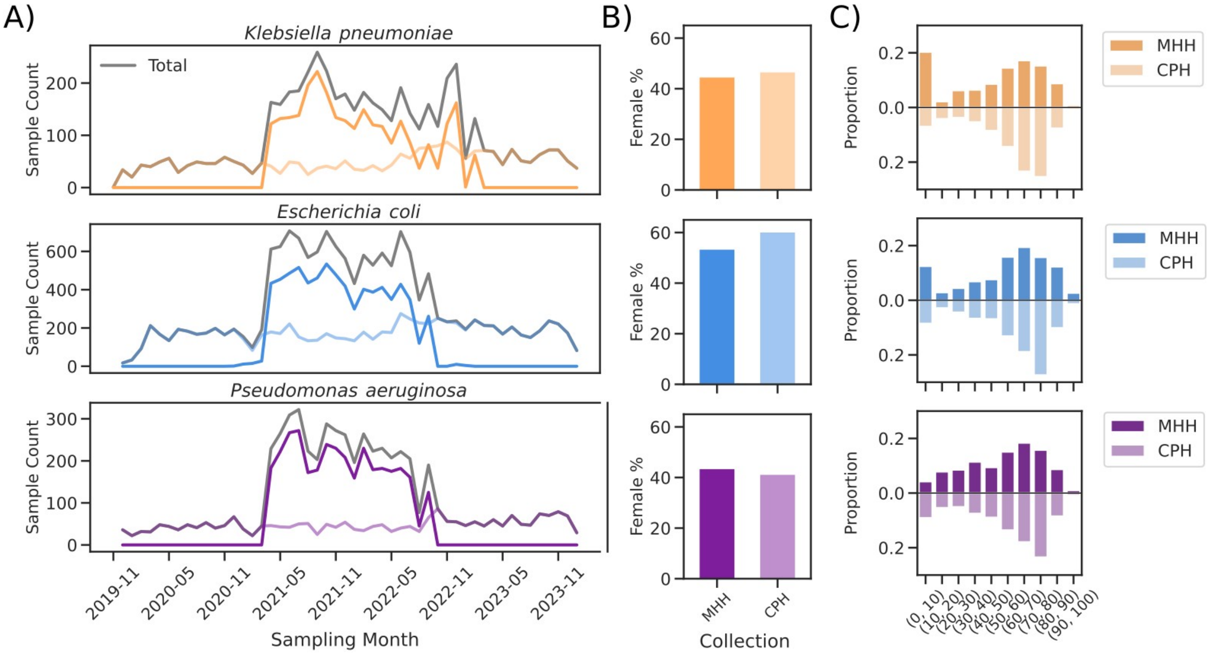


**Supplementary Figure 1. Sample distribution of the whole-genome sequencing surveillance dataset. A)** Sample counts per month and hospital. **B)** Percentage of female patients across species and hospital settings. **C)** Proportion of patients within age ranges across species and hospital settings.

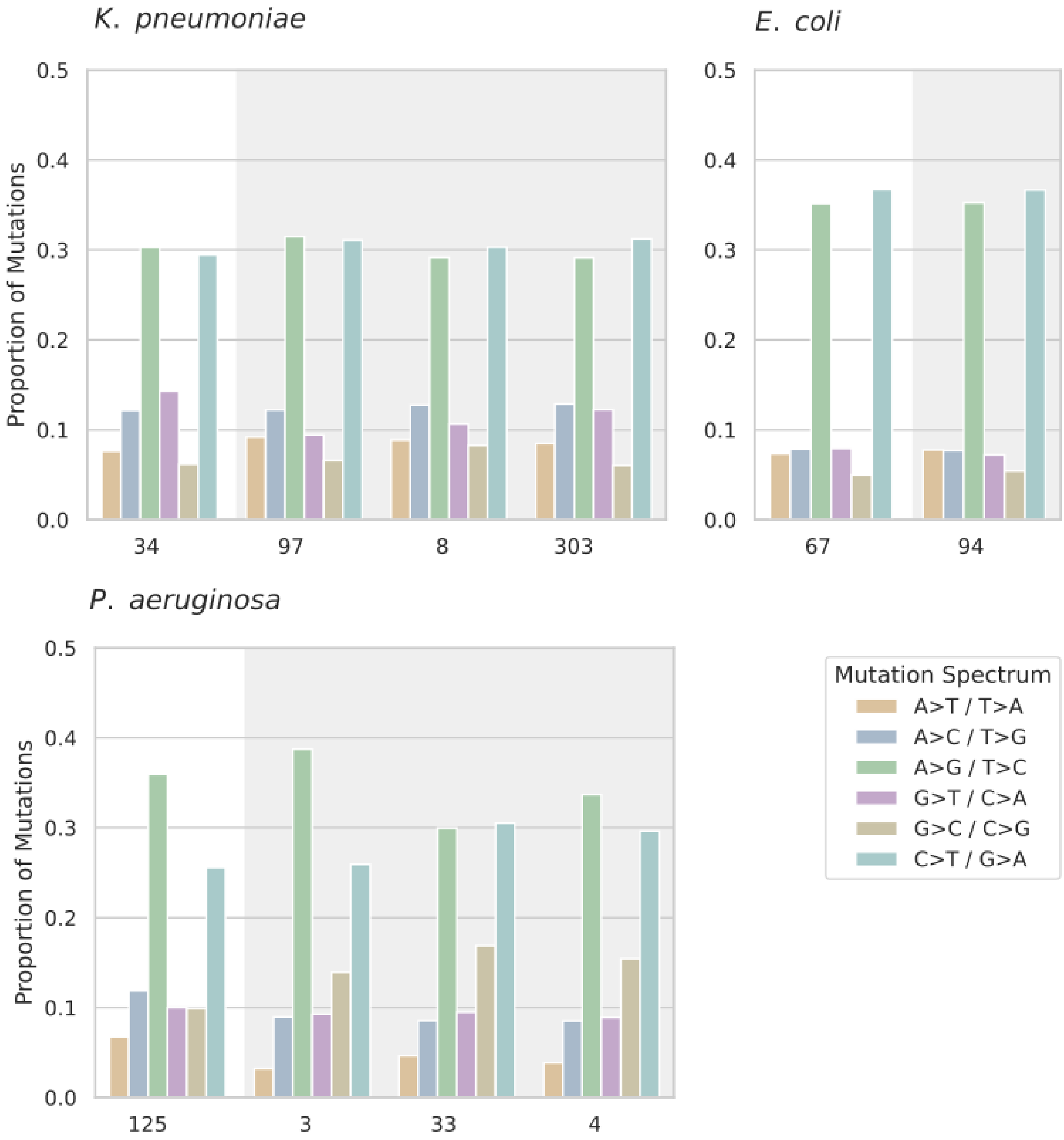


**Supplementary Figure 2. Mutation spectra across bacterial isolates.** Proportion of SNPs types observed in isolates of *K. pneumoniae*, *E. coli* and *P. aeruginosa*. Shaded backgrounds distinguish isolates with a normal mutation rate (white) compared to hypermutator strains (grey).

**Supplementary Figure 3. Comparison of the presence of an epidemiological link and SNP distribution in predicted transmission events with the time-aware threshold (180d) and the fixed SNP threshold (20SNP).** The left panels show confusion matrices of predicted transmission (T=1) versus no transmission (T=0) for isolate-pairs with an epidemiological link (Epi Link) or not (No Epi Link). The right panels show the distribution of SNP distances under the different settings

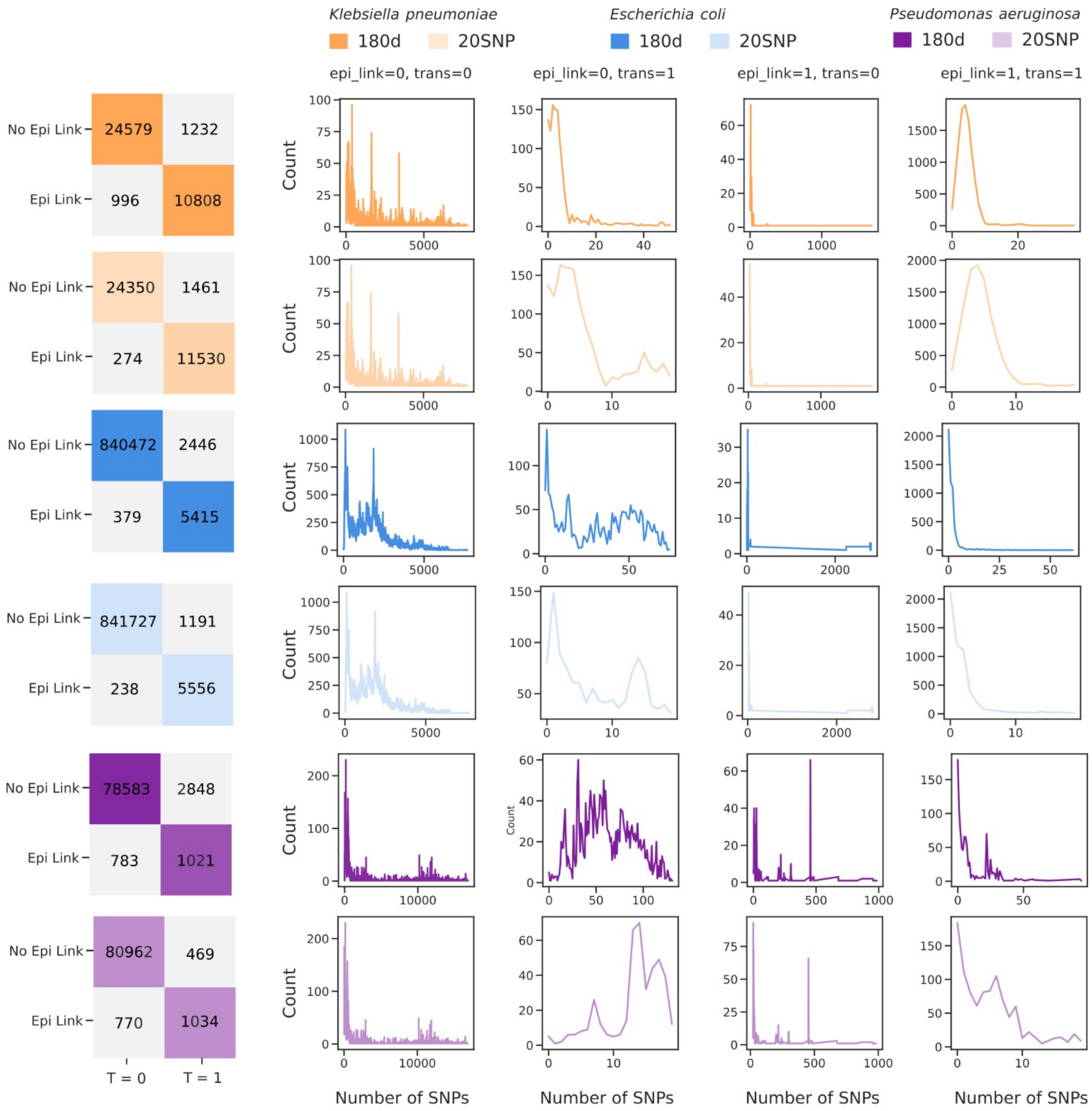


(combination of epi links and predicted transmission). Results are shown for the three species with colors distinguishing between the two different thresholds (180d and 20SNPs).

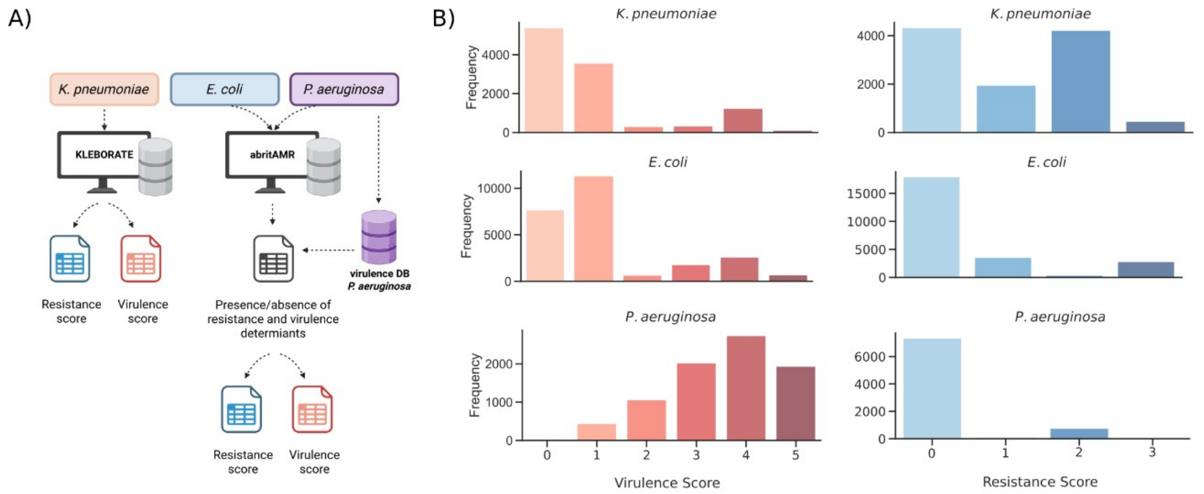


**Supplementary Figure 4. Virulence and resistance scores prediction. A)** Workflow for the generation of the virulence and resistance scores. **B)** Frequency distribution of the virulence and resistance score for *K. pneumoniae*, *E. coli* and *P. aeruginosa*.

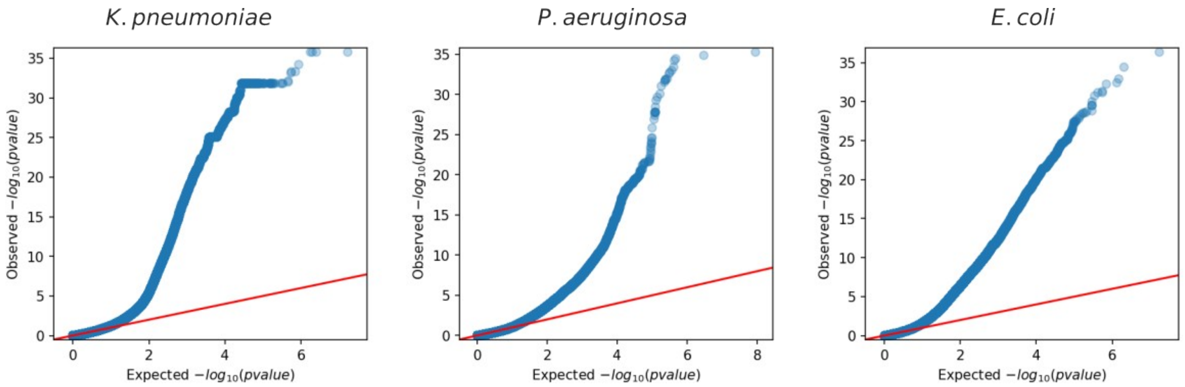


**Supplementary Figure 5. QQ-plots from the unitigs association for *K. pneumoniae, P. aeruginosa* and *E. coli*.**

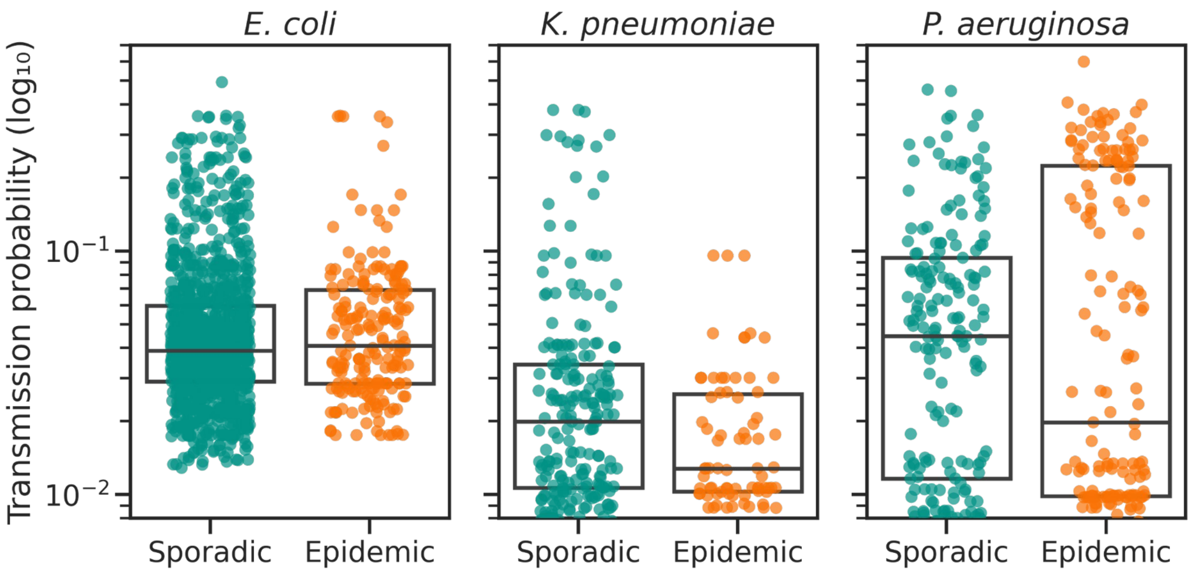


**Supplementary Figure 6. Model performance in predicting the transmissibility potential of new isolates.** Predictions from the ridge regression model applied to genomes collected in 2024 show no significant difference in predicted transmission probability between sporadic and epidemic strains across the three species.
